# Longitudinal association of remnant cholesterol with cognitive decline vary by lipid-lowering therapy: a population-based cohort study

**DOI:** 10.1101/2024.11.16.24317444

**Authors:** Jianian Hua, Jianye Dong, Ying Chen, Haibin Li, Qingmei Chen

## Abstract

**Objective:** Although the association between remnant cholesterol (RC) and cognitive impairment has been reported, the association of RC with cognitive decline remains scarce. Also, the role of lipid-lowering therapy in the association is unclear. The study aimed to examine the longitudinal associations of RC with cognitive decline by lipid-lowering drug use status.

**Methods:** The study utilized data from wave 2 (2004-2005) to wave 8 (2016-2017) of the English Longitudinal Study of Ageing (ELSA). Global cognitive functions at baseline (wave 2) and during the follow-up (waves 3-8) were assessed by integrating three cognitive domains, including memory capacity, semantic fluency, and orientation. Multivariate-adjusted linear mixed models were employed to examine the longitudinal associations, with results presented as *β* [95% confidence interval (CI)] in standard deviation (SD)/year.

**Results:** Of the 5053 participants ultimately included, 55.4% were female and the mean age (SD) was 65.7 (9.3) years. Per 1 mmol/L increment in RC was significantly associated with a faster rate of cognitive decline (*β* = −0.010 SD/year, 95% CI: -0.019, -0.001). Furthermore, we observed that the association pattern between RC and cognitive decline only in the non-lipid-lowering drug group (*β* = -0.019 SD/year, 95% CI: -0.031, -0.007), but not in the lipid-lowering drug group (*β* = 0.007 SD/year, 95% CI: -0.006, 0.020), with a significant interaction (*P* = 0.015). Similar findings were observed for the three cognitive domains.

**Conclusions:** Higher baseline RC levels were associated with steeper cognitive decline. Regular use of lipid-lowering drugs during follow-up might attenuate the accelerated cognitive decline caused by high RC.

## Introduction

Aging is often accompanied by varying levels of cognitive decline, which can, in severe cases, lead to dementia^1^. It is estimated that dementia cases globally will exceed 150 million by 2050^2^, placing a considerable strain on families and society. In the absence of effective dementia treatments, studies have demonstrated that targeting modifiable risk factors could decrease dementia incidence by as much as 40%^3^. Additionally, accelerated cognitive decline represents a rapidly growing public health burden and is the key clinical characteristic and the early symptom of dementia^4^. Therefore, identifying modifiable risk factors for cognitive decline could not only enhance quality of life but also potentially prevent dementia.

Numerous epidemiologic studies have reported abnormal lipid levels as key risk factors for the development of cognitive impairment or dementia^5–9^. For instance, a cohort study with over 20 years of follow-up confirmed that elevated total cholesterol (TC) and low-density lipoprotein cholesterol (LDL-C) in midlife were significantly associated with cognitive decline in later life^8^. A prospective study based on 18,668 older adults reported that higher high-density lipoprotein cholesterol (HDL-C) levels were associated with an elevated risk of dementia^9^. However, previous studies have focused primarily on traditional lipid indicators such as TC, HDL-C, and LDL-C. As a novel lipid indicator, remnant cholesterol (RC) has attracted considerable interest due to its robust predictive capacity^10,11^. RC is the cholesterol found in all triglyceride-rich lipoproteins, which includes all cholesterol in the body apart from HDL-C and LDL-C^12^. Existing evidence has indicated that RC could predict cardiovascular disease as effectively as, or even more reliably than, other lipid markers^10,11^. Previous studies have reported associations between RC and cognitive impairment or dementia ^13–17^. However, knowledge about the association between RC levels and cognitive decline remains scarce. Hence, exploring the longitudinal association between RC and cognitive decline is urgent. Notably, while lipid-lowering medications were associated with a decreased risk of cognitive impairment and dementia^18,19^, it is unknown whether they can mitigate the long-term effects of elevated RC on cognitive decline.

Drawing on repeated-measures data from the English Longitudinal Study of Ageing (ELSA) cohort, the present study sought to investigate the longitudinal association between RC and cognitive decline and to examine further whether this association varied among participants taking or not taking lipid-lowering drugs.

## Methods

### Study population

This study utilized data from the ELSA cohort, with details on its objectives, design, and methods described previously^20^. In short, ELSA is a prospective, nationally representative aging cohort that collects socioeconomic, physical and mental health, cognitive, biological, and genetic data from older residents (≥ 50 years) in England. The ELSA was initiated in 2002-2003 (Wave 1) and participants were followed up every two years until 2018-2019 (Wave 9). Due to the absence of lipid measurements in Wave 1 and medication information in Wave 9, data from Wave 2 (2004-2005) to Wave 8 (2016-2017) were utilized for this study. Therefore, the Wave 2 in this study was considered as the baseline. Of the 9,432 individuals who participated in Wave 2 of the ELSA, 163 were excluded from the study for lacking cognitive assessment data. Furthermore, we excluded participants based on the following criteria: (1) age below 50 years (*n*=252), (2) diagnosis of Parkinson’s disease, Alzheimer’s disease, or dementia (*n*=103), (3) missing RC data (*n*=3,248), and (4) missing covariate information (*n*=42). Also, we excluded 571 participants who were lost to follow-up from Wave 3 to Wave 8. Finally, 5,053 participants with complete baseline data and at least one cognitive evaluation from Wave 3 to Wave 8 were included in the current analysis. A flowchart outlining the selection process of study participants is displayed in **Figure 1**. Ethical approval for the ELSA was obtained from the London Multicentre Research Ethics Committee (MREC/01/2/91), and informed consent was gathered from all participants.

**Figure 1.**
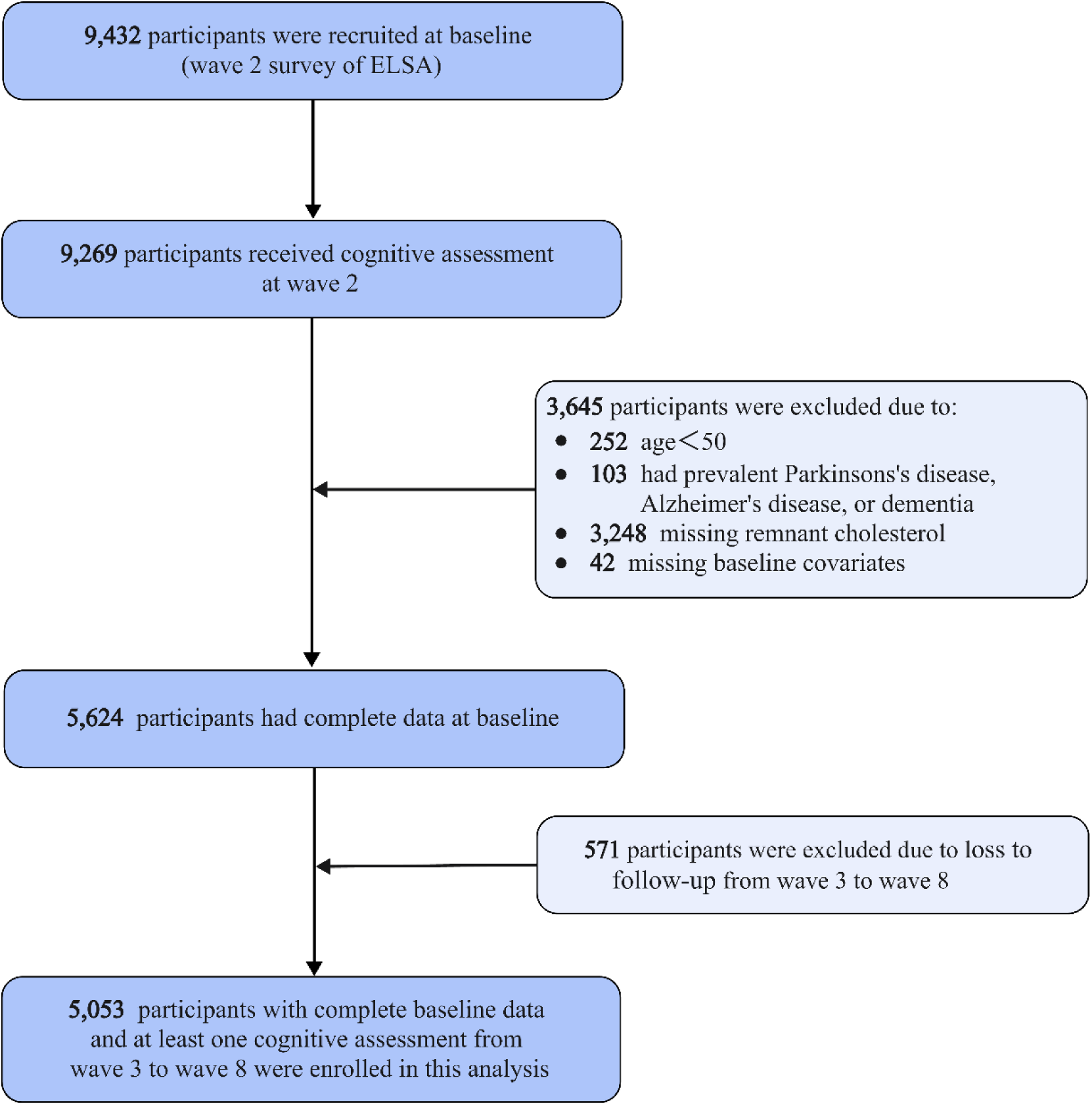
Flowchart of study participant selection. Abbreviations: ELSA, English Longitudinal Study of Ageing.

### Measurement of RC and lipid-lowering therapy

In the Wave 2 surveys of the ELSA, blood samples were collected by trained nurses and forwarded to the Royal Victoria Hospital in Newcastle, UK for a variety of laboratory analyses including lipids^21^. Detailed information regarding the analysis of blood samples is provided in a previous document^22^. Briefly, in the Wave 2 of ELSA, about two-thirds (64.1%) of the blood samples were taken in the fasting state. However, according to the current consensus, no clinically significant differences between fasting and non-fasting lipid levels are observed^23,24^. Therefore, we analyzed lipids regardless of fasting status. LDL-C was derived using the Friedewald formula in ELSA^25^. Based on previous studies^10,17^, the RC concentration was approximated as the TC concentration minus the LDL-C concentration and subsequently the HDL-C concentration. Lipid-lowering therapy was measured by the question “Whether taking medication to lower cholesterol level or prevent level becoming high”. Information about the use of lipid-lowering drugs was interviewed in Waves 3 to 8 of the ELSA.

### Assessment of cognitive function

Three cognitive domains were used to evaluate the cognitive performance of the participants in each wave, including memory performance, semantic fluency tests, and orientation ability. Memory performance was assessed using immediate and delayed recall tasks, where participants were asked to recall 10 unrelated words. One point was given for each correctly recalled word, with a maximum possible score of 20, where higher scores indicated better memory performance. Participants in the semantic fluency test were instructed to name as many animals as possible in one minute, excluding repetitions and non-animal words. The score range was from 0 to 50, with higher scores representing better semantic fluency. At baseline, the median score for semantic fluency was 20, with an interquartile range of 16 to 24. Orientation ability was assessed by inquiring about the year, month, day, and day of the week, with one point awarded for each correct answer, yielding a score from 0 to 4. The validity and reliability of these three cognitive tests have been extensively validated and reported in previous studies^26,27^. Also, to facilitate the comparison of effect sizes across various cognitive tests, we applied a *z*-transformation to the cognition score of each individual (*i*) at each time point (*t*), following the formula below: *z*-scoreit = (*scoreit* − *mean scorebaseline*) / *SDbaseline*. A *z*-value of -1 at any time point signified that the individual’s cognitive performance was 1 SD below the mean cognitive score observed at baseline ^26,27^. To provide a summary of the three cognitive domains, we averaged the *z*-values of the three tests and subsequently standardized the average to the baseline values in the same way to generate a global cognitive *z*-score.

### Covariates

We adjusted for covariates associated with RC or cognitive function, which included age, sex, education level [≥ level 3 National Vocational Qualification (NVQ3)/General Certificate of Education (GCE) A level vs. < NVQ3/GCE A level], marital status (married vs. unmarried), current smoking or drinking (yes vs. no), body mass index (BMI), physical activity (inactive vs. moderate-vigorous activity), depressive symptoms (yes vs. no), LDL-C, hypertension (yes vs. no), diabetes (yes vs. no), and stroke (yes vs. no) ^14,26,27^. Details of covariates are provided in **Supplemental Methods**.

### Statistical analyses

Descriptive statistics were performed for continuous variables [expressed as mean (SD)] and categorical variables [presented as n (%)] to depict the distribution of baseline characteristics of participants who used lipid-lowering drugs during the follow-up period (lipid-lowering group) and those who did not (non-lipid-lowering group). Analysis of variance (ANOVA) or chi-square tests were conducted to compare the differences in characteristics between the two groups.

To examine the dose-response relationship between baseline RC and the rate of global cognitive decline, restricted cubic spline (RCS) models with ordinary least squares regression were fitted^28^. The optimal number of spline knots was determined according to the Akaike Information Criterion, and ANOVA was applied to test the nonlinear terms of the model effects^28^. The rate of cognitive decline was calculated by subtracting the cognitive *z*-score at the last follow-up visit from the *z*-score at the first follow-up visit and dividing it by the follow-up duration (years) for each participant^28^. Since data from multiple repeated cognitive assessments were available, we constructed a linear mixed model (LMM) to examine the longitudinal association between continuous RC and the rate of cognitive decline. The LMM takes into account correlations between repeated cognitive assessments within the same individual, variability in baseline cognitive scores, inter-individual differences in the rate of cognitive decline, and missing cognitive assessments^26,29^. Intercept and slope (time) were fitted as random effects in all models. Fixed effects were fitted for intercept, time, RC, RC × time interaction, and covariates. The interaction term reflected the average difference in change rates of cognitive *z* scores over the follow-up. Two LMMs adjusting for different covariates in the fixed effects were constructed: Model 1 adjusted for age and sex; Model 2 further adjusted for baseline LDL-C, lipid-lowering therapy, BMI, education, marital status, physical activity, current smoking and drinking status, depressive symptoms, hypertension, diabetes, and stroke plus Model 1. Moreover, the analyses were repeated for the lipid-lowering and non-lipid-lowering groups, seperately. To investigate whether lipid-lowering therapy modified the association of RC with cognitive decline, we constructed another LMM with fixed effects including intercept, time, RC × time, lipid-lowering therapy, lipid-lowering therapy × time, lipid-lowering therapy × RC × time, and all covariates. The ‘lipid-lowering therapy × RC × time’ interaction term reflected the modification effect of lipid-lowering drugs on the association between RC and cognitive decline. To visualize the role of taking lipid-lowering drugs, we fitted cognitive trajectories for four participants at RC concentrations of 0.5 mmol/L and 2.0 mmol/L and with or without lipid-lowering drugs.

Several additional analyses were performed to evaluate the robustness of our findings. First, we performed stratification analyses by age (<60 years/≥ 65 years), gender (male/female), and educational level (< NVQ3/GCE A level/≥NVQ3/GCE A level) to test whether these covariates modified the effect of RC on rate of cognitive decline by using interaction terms. Second, we re-performed the association analyses after excluding participants who experienced a stroke during the follow-up, as stroke has been repeatedly associated with accelerated cognitive decline^30^. In addition, we reanalyzed the relationship between baseline RC levels and rates of cognitive decline after restricting participants to those who took all waves of the cognitive assessment.

Statistical analyses were performed with SAS 9.4 (SAS Institute, Cary, NC, USA) and R Statistical Software (version 4.4). Statistical significance was considered when the two-tailed *P* value was below 0.05.

## Results

### Baseline characteristics

The distribution of baseline characteristics among the study participants is summarized in **Table 1**. Of the 5,053 participants included in the study, 55.4% were female, and the mean age was 65.7 years, with an SD of 9.3 years. During the follow-up period, 1889 (37.4%) participants reported taking lipid-lowering medications. The mean RC concentration for all participants was 0.8 mmol/L (SD = 0.4), which was significantly higher in the group taking lipid-lowering medications (0.9 mmol/L) than in the non-medication group (0.7 mmol/L). Additionally, participants in the medicated group were more likely to have lower education levels, be physically inactive, have a higher BMI, and exhibit a higher prevalence of hypertension, diabetes, and stroke compared to the non-medicated group (*P* < 0.05).

**Table 1.**
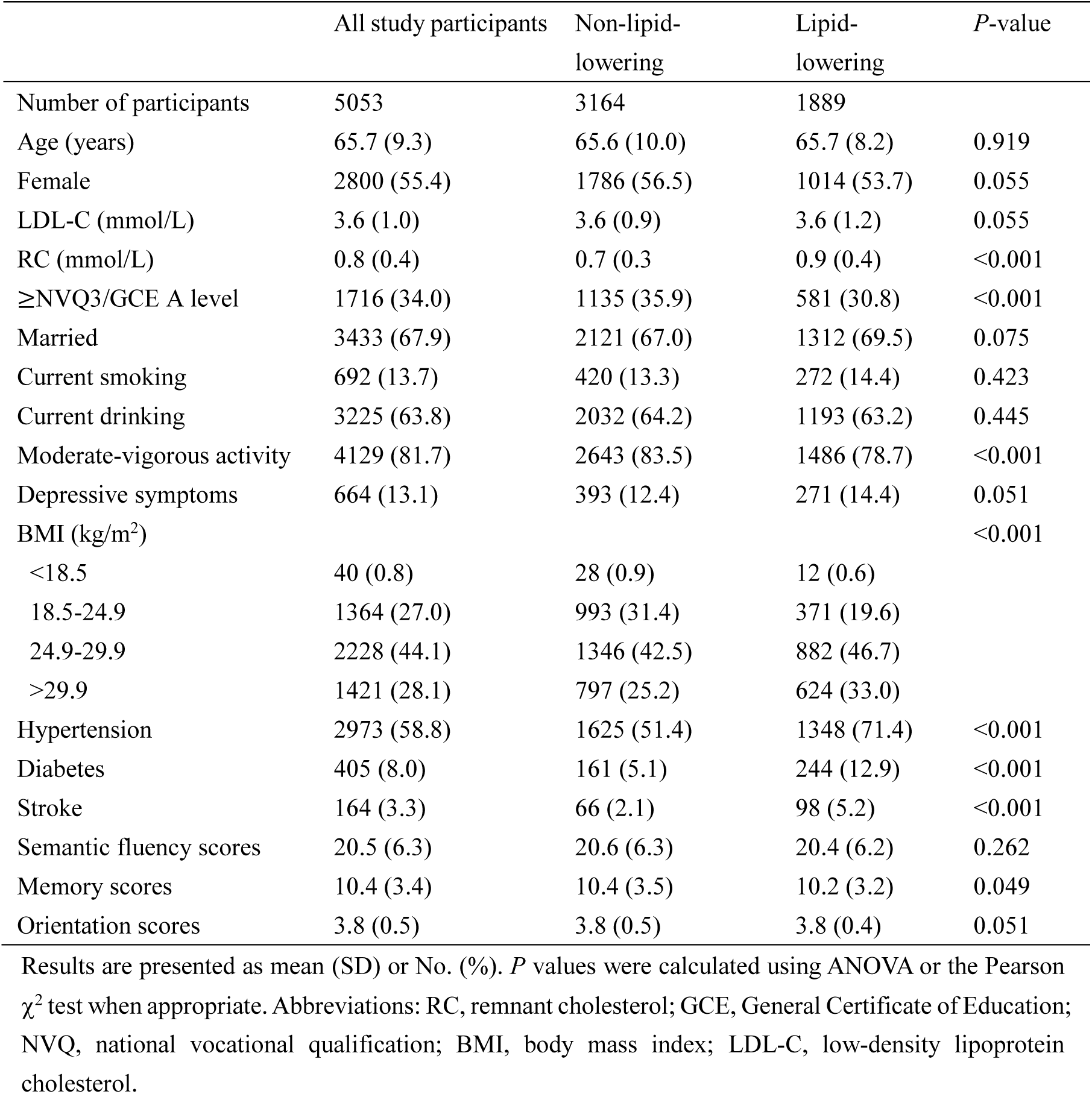
Baseline characteristics of study participants according to lipid-lowering drug use during the follow-up. Results are presented as mean (SD) or No. (%). *P* values were calculated using ANOVA or the Pearson χ^2^ test when appropriate. Abbreviations: RC, remnant cholesterol; GCE, General Certificate of Education; NVQ, national vocational qualification; BMI, body mass index; LDL-C, low-density lipoprotein cholesterol.

|  | All study participants | Non-lipid-lowering | Lipid-lowering | *P*-value |
| --- | --- | --- | --- | --- |
| Number of participants | 5053 | 3164 | 1889 |  |
| Age (years) | 65.7 (9.3) | 65.6 (10.0) | 65.7 (8.2) | 0.919 |
| Female | 2800 (55.4) | 1786 (56.5) | 1014 (53.7) | 0.055 |
| LDL-C (mmol/L) | 3.6 (1.0) | 3.6 (0.9) | 3.6 (1.2) | 0.055 |
| RC (mmol/L) | 0.8 (0.4) | 0.7 (0.3) | 0.9 (0.4) | <0.001 |
| ≥NVQ3/GCE A level | 1716 (34.0) | 1135 (35.9) | 581 (30.8) | <0.001 |
| Married | 3433 (67.9) | 2121 (67.0) | 1312 (69.5) | 0.075 |
| Current smoking | 692 (13.7) | 420 (13.3) | 272 (14.4) | 0.423 |
| Current drinking | 3225 (63.8) | 2032 (64.2) | 1193 (63.2) | 0.445 |
| Moderate-vigorous activity | 4129 (81.7) | 2643 (83.5) | 1486 (78.7) | <0.001 |
| Depressive symptoms | 664 (13.1) | 393 (12.4) | 271 (14.4) | 0.051 |
| BMI (kg/m<sup>2</sup>) |  |  |  | <0.001 |
| <18.5 | 40 (0.8) | 28 (0.9) | 12 (0.6) |  |
| 18.5-24.9 | 1364 (27.0) | 993 (31.4) | 371 (19.6) |  |
| 24.9-29.9 | 2228 (44.1) | 1346 (42.5) | 882 (46.7) |  |
| >29.9 | 1421 (28.1) | 797 (25.2) | 624 (33.0) |  |
| Hypertension | 2973 (58.8) | 1625 (51.4) | 1348 (71.4) | <0.001 |
| Diabetes | 405 (8.0) | 161 (5.1) | 244 (12.9) | <0.001 |
| Stroke | 164 (3.3) | 66 (2.1) | 98 (5.2) | <0.001 |
| Semantic fluency scores | 20.5 (6.3) | 20.6 (6.3) | 20.4 (6.2) | 0.262 |
| Memory scores | 10.4 (3.4) | 10.4 (3.5) | 10.2 (3.2) | 0.049 |
| Orientation scores | 3.8 (0.5) | 3.8 (0.5) | 3.8 (0.4) | 0.051 |

Results are presented as mean (SD) or No. (%). *P* values were calculated using ANOVA or the Pearson $\chi^2$ test when appropriate. Abbreviations: RC, remnant cholesterol; GCE, General Certificate of Education; NVQ, national vocational qualification; BMI, body mass index; LDL-C, low-density lipoprotein cholesterol.

### Association between baseline RC bevels and rate of cognitive decline

The RCS analysis did not provide evidence of a nonlinear relationship between baseline RC levels and the rate of cognitive decline (**Figure S1**, *P*nonlinear = 0.737). **Table 2** outlines the associations between baseline RC levels and the rate of change in cognitive scores. In the multivariable-adjusted model, each 1 mmol/L increase in RC was remarkably associated with a greater rate of cognitive decline, as indicated by a reduction of -0.010 SD/year (95% CI: -0.019, -0.001) in global cognitive *z*-scores. Furthermore, we observed similar decline trends in the cognitive subdomains of semantic fluency [*β =* -0.007 SD/year (95% CI: -0.015, 0.000)], memory capacity [*β =* -0.006 SD/year (95% CI: -0.013, 0.001)], and orientation ability [*β =* -0.004 SD/year (95% CI: -0.014, 0.007)], although the associations did not reach statistical significance.

**Table 2.**
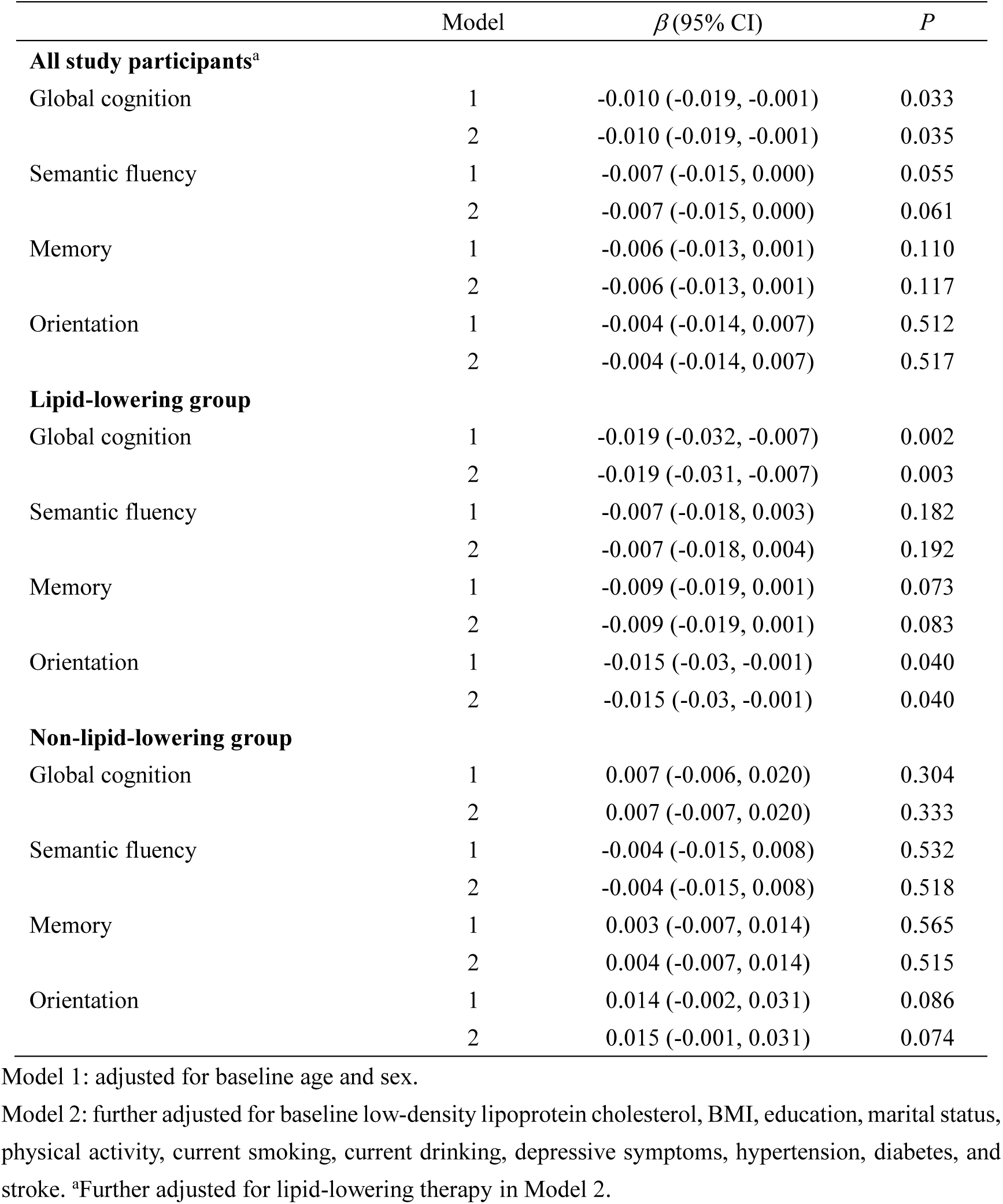
The association between remnant cholesterol level at baseline and decline rates of cognitive *z* scores (SD/year) by lipid-lowering drug use during the follow-up.

### Role of lipid-lowering drug use in the association

Figure 2 demonstrates the dose-response associations between baseline RC levels and rates of global cognitive decline stratified by lipid-lowering drug use or not. Specifically, in the non-drug group, baseline RC levels were linearly associated with accelerated rates of cognitive decline (*P*nonlinear = 0.849). In contrast, in the drug-using group, we observed a slower rate of cognitive decline with increasing RC levels (*P*nonlinear = 0.111). The findings of further stratification analyses by lipid-lowering drug use are presented in **Table 2**. After adjusting for age, sex, LDL-C, BMI, education, marital status, physical activity, current smoking and drinking status, depressive symptoms, hypertension, diabetes, and stroke, we detected a significant association of higher baseline RC concentrations with a faster rate of cognitive decline only in the non-drug group [global cognition: *β =* -0.019 SD/year (95% CI: -0.031, -0.007)] but not in the drug group [global cognition: *β =* 0.007 SD/year (95% CI: -0.006, 0.020)], with a significant interaction (*P* for interaction = 0.015). Also, similar findings were observed in the cognitive subdomains of memory capacity [non-lipid-lowering group vs. lipid-lowering group: -0.009 (-0.019, 0.001) vs. 0.004 (-0.007, 0.014)] and orientation [non-lipid-lowering group vs. lipid-lowering group: -0.015 (-0.030, - 0.001) vs. 0.015 (-0.001, 0.031)]. As illustrated in Figure 3, the global cognitive scores of all four participants gradually declined over time. Among the two participants not taking lipid-lowering drugs, the one with higher RC showed a steeper rate of decline. Conversely, among the two participants who took lipid-lowering drugs, the participants with higher RC exhibited a slower decline rate. The effect of higher RC levels on the rate of cognitive decline was not as pronounced in the lipid-lowering group as in the non-lipid-lowering group. Our results suggested that the use of lipid-lowering drugs might attenuate the adverse impact of elevated RC on cognitive decline.

**Figure 2.**
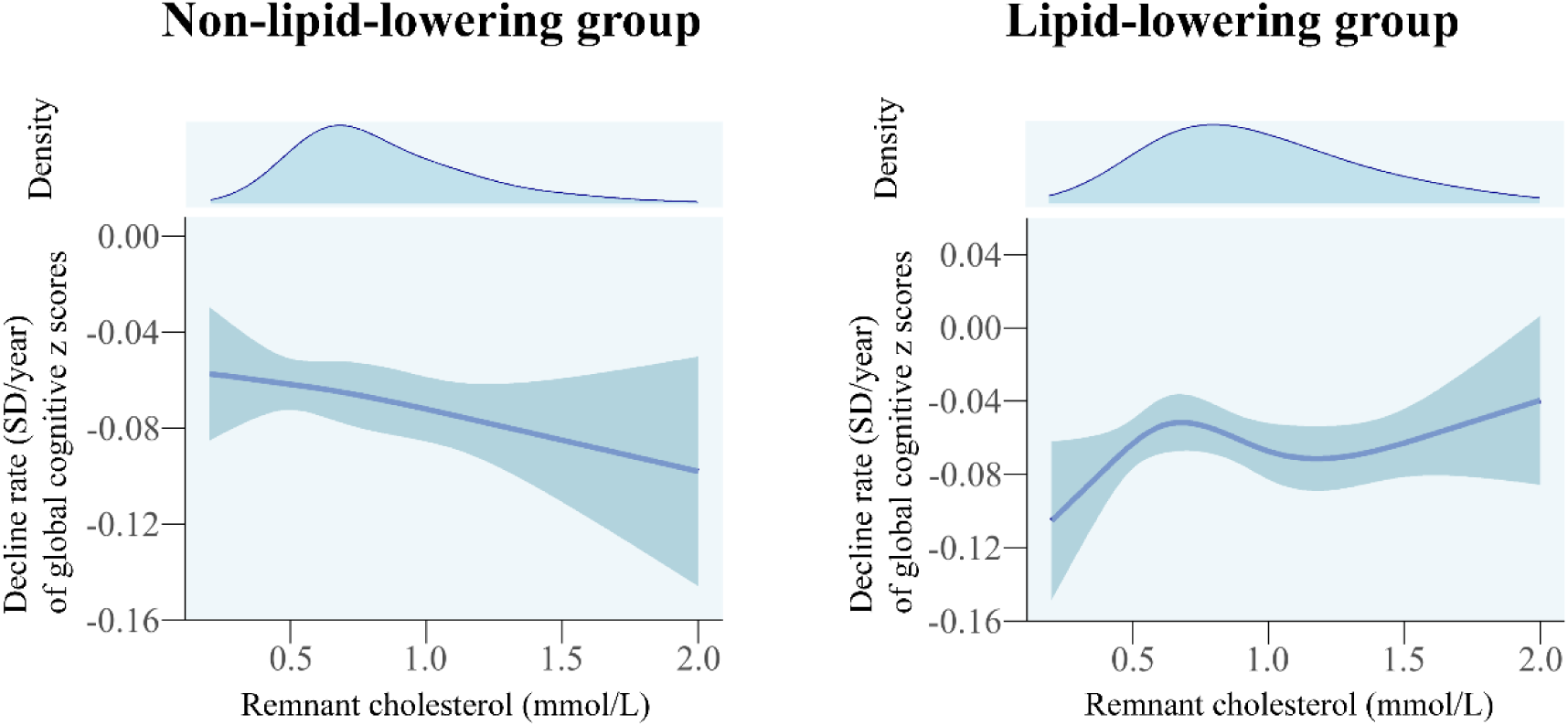
Dose-response associations of remnant cholesterol levels at baseline with rate of cognitive decline over the follow-up according to lipid-lowering drug use. Three knots were placed at the 10th, 50th, and 90th percentiles of remnant cholesterol values for the non-lipid-lowering group, while four knots (5th, 35th, 65th, and 95th) were placed for the lipid-lowering group. Adjusted for age, sex, low-density lipoprotein cholesterol, body mass index, education, marital status, physical activity, current smoking, current drinking, depressive symptoms, hypertension, diabetes, and stroke.

**Figure 3.**
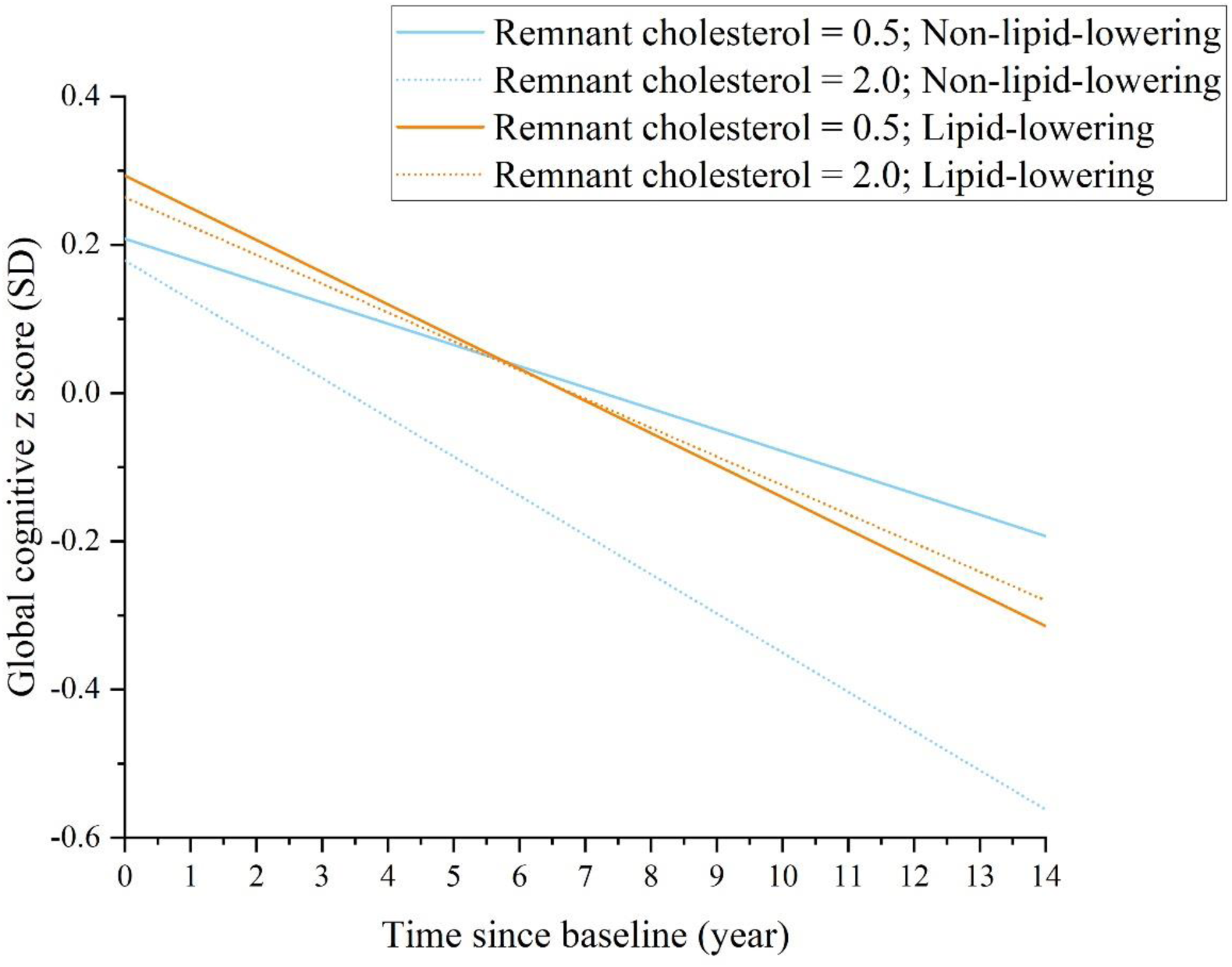
Trajectories of cognitive *z* scores according to remnant cholesterol levels and lipid-lowering therapy. Baseline covariates were set to the most common characteristics: 65 years old, female, LDL-C of 3.6 mmol/L, education level below NVQ3/GCE A level, married, current drinker but non-smoker, BMI between 24.9 and 29.9, engaging in moderate-to-vigorous activities, non-depressed, and free from hypertension, diabetes, or stroke. Random effects were set to 0. The blue solid line indicates a participant with a remnant cholesterol level of 0.5 mmol/L who had not used lipid-lowering drugs during the follow-up; the blue dotted lines, a remnant cholesterol level of 2.0 mmol/L and had not used lipid-lowering drugs; the yellow solid line, a remnant cholesterol level of 0.5 mmol/L and used lipid-lowering drugs during the follow-up; the yellow dotted line, a remnant cholesterol level of 2.0 mmol/L and used lipid-lowering drugs. Adjusted for baseline LDL-C, BMI, education, marital status, physical activity, current smoking, current drinking, depressive symptoms, hypertension, diabetes, and stroke. Abbreviations: LDL-C, low-density lipoprotein cholesterol; BMI, body mass index; GCE, General Certificate of Education; NVQ, national vocational qualification.

### Sensitivity and subgroup analyses

Subgroup differences were detected in the associations between RC and the rate of cognitive decline by gender. Specifically, the detrimental impact of high RC on cognitive decline was more pronounced in females than in males (Global cognition: *P* for interaction = 0.009; Memory performance: *P* for interaction = 0.017; **Table S1**). Moreover, the associations were similar in subgroups of age and education levels (*P* for interaction > 0.05, **Tables S2 and S3**). Sensitivity analyses excluding participants who experienced stroke during follow-up yielded similar findings to the primary analyses (**Table S4**). The repeated analyses with the participants restricted to those who took all cognitive assessments also did not change the robustness of the associations (**Table S5**).

## Discussion

Drawing on a large prospective cohort with more than 12 years of follow-up, this study revealed that higher baseline RC levels were significantly associated with steeper subsequent cognitive decline among adults aged 50 years and older in England. Moreover, we identified a significant modifying effect of lipid-lowering drug use on the association. Specifically, we found that the adverse effects of high levels of RC on cognitive decline was observed only in participants not taking lipid-lowering drugs, whereas such effects were absent in participants taking lipid-lowering drugs.

To our knowledge, the present study is the first to examine the longitudinal association between higher RC concentration at baseline and faster cognitive decline using data from seven waves of repeated measures. Our findings reinforced and expanded upon previous cross-sectional studies on this topic^13–16^. A case-control study involving 36 mild cognitive impairment (MCI) cases and 38 healthy controls reported a positive association between plasma RC levels and the risk of MCI^13^. Another case-control study from Wuhan, China, involving 1,007 community-based older participants, also observed a significant relationship between RC levels and MCI^15^. Besides, in a cross-sectional survey of 1,331 older American participants from the National Health and Nutrition Examination Survey (NHANES), Liu et al. observed that elevated RC levels were linked to a higher probability of cognitive impairment^14^. Similarly, another NHANES-based study found cross-sectional relationships between RC levels and cognitive domains such as verbal fluency and memory capacity^16^. However, these studies were cross-sectional and mainly focused on dichotomous cognitive impairments. The categorization of continuous variables can result in a loss of valuable information, underscoring the necessity of longitudinal designs to explore the associations between RC concentrations and continuous cognitive performance. Notably, two prospective cohort studies have reported independent associations between higher RC levels and increased risk of dementia or cognitive impairment, providing some longitudinal evidence in support of our findings^17,31^. However, both studies measured cognitive function at a single time point and did not consider the temporal variability in cognitive performance^17,31^. Using longitudinal repeated-measures data with over 12 years of follow-up, our study provided first-hand evidence that higher baseline RC levels are associated with faster rates of cognitive decline. By using multi-wave data, the study captured changes in cognitive ability over time and accounted for individual differences, increasing the sensitivity of detecting the effect of baseline RC levels on cognitive decline. Notably, we observed that women were more vulnerable to the cognitive decline associated with elevated RC, similar to previous findings^16,32^. One possible explanation is that women will undergo a sharp loss of estradiol after menopause thereby decreasing the protective effect on the nervous system^32^. Our findings emphasized the relevance of early monitoring of RC levels to identify individuals at higher risk of cognitive decline, especially for females.

Although the precise mechanism by which high RC levels contribute to cognitive decline is not yet known, several potential explanations have been suggested. First, a study using Mendelian randomization (MR) design found poor lipid profiles in *APOE* ɛ4 gene carriers who were significantly associated with cognitive impairment^31^. Second, RCs smaller than 70 nanometers can cross the walls of arteries, potentially contributing to atherosclerosis within the brain and reducing cerebral blood flow, which may impair cognitive function^33,34^. Third, elevated RC levels are associated with impaired vascular endothelial function, which can increase the secretion of β-amyloid, thereby potentially increasing the risk of neurodegeneration^35,36^. In addition, elevated RC levels may worsen oxidative stress and pro-inflammatory responses in vascular endothelial cells, compromising the blood-brain barrier and ultimately contributing to cognitive decline^37–39^. Additional biological mechanisms by which elevated RC levels may contribute to cognitive decline include reduced antithrombotic capacity and dysregulated immune function^39^. However, further randomized clinical trials are necessary to confirm these potential biological mechanisms.

The findings from our stratification analysis demonstrated that taking lipid-lowering drugs could attenuate the adverse impact of high RC on accelerated cognitive decline. Specifically, in the non-medicated group, higher baseline RC levels were associated with more rapid cognitive decline. In contrast, in the medicated group, although cognitive decline was observed over time, higher baseline RC levels were associated with a slower rate of decline. Our findings underscored for the first time the benefits of regular lipid-lowering drugs in high-RC patients in slowing the rate of cognitive decline and thus preventing or delaying the incidence of dementia. However, the role of lipid-lowering drugs in preventing cognitive impairment or dementia has been debated. Several studies have reported the beneficial role of lipid-lowering drugs in preventing cognitive decline and dementia^40,41^. For example, a longitudinal cohort study from Sweden found that regular statin use was associated with improved cognitive function compared to non-drug users^40^. Still, some studies have indicated that taking lipid-lowering drugs was not statistically associated with cognitive function^42–44^. A review of eleven randomized trials concluded that lipid-lowering drugs had no significant effect on cognitive performance^42^, which might be attributable to the relatively short observation period. One possible explanation is that the benefits of lipid-lowering drugs on cognitive function might require a longer follow-up period to fully emerge. In support of this, the present study observed a significant benefit of lipid-lowering drugs in delaying cognitive decline in a longitudinal dataset with over 12 years of follow-up. A systematic review and meta-analysis that included 27 qualitative and quantitative studies similarly concluded that data from long-term follow-up may be supportive of the beneficial role of statins in dementia prevention^45^. Well-designed studies with longer-term follow-up are warranted to confirm the benefits of lipid-lowering medications in slowing cognitive decline.

A notable strength of the study is that it is the first to explore the relationship between baseline RC levels and the rate of cognitive decline and the role of lipid-lowering drug use based on a nationally representative cohort over a long 12-year follow-up. Another key advantage is that multiple repeated measures of cognitive function allowed us to capture the long-term trajectory of cognitive decline. Moreover, several limitations should be mentioned. First, although we have adjusted for multiple covariates, the possibility of unmeasured and residual confounders remains, such as the *APOE* genotype (genetic information not available for ELSA). Second, the nature of observational research makes it impossible to rule out the possibility of reverse causation. Future large-scale well-designed clinical trials or MR studies are required to confirm the causality of the observed association. Third, the study population was limited to those aged 50 and over in England, which may restrict the extrapolation of the findings to other ethnic and age groups. Finally, the RC in the study was calculated indirectly from TC, HDL-C, and LDL-C values instead of being directly measured. Nonetheless, it is important to note that the reliability of the calculated RC has been validated in previous studies^10,17,46^. Additionally, a strong correlation has been established between the RC concentrations derived from both methods^47^. More importantly, compared to direct measurements, calculating methods have practical advantages in large-scale population studies.

## Conclusion

The longitudinal study indicated that elevated baseline RC levels were associated with accelerated cognitive decline. Furthermore, our findings demonstrated that taking lipid-lowering drugs might mitigate the adverse impact of the accelerated cognitive decline caused by high RC. Our findings provided evidence for early monitoring of RC levels to slow the rate of cognitive decline and emphasized the benefits of regular lipid-lowering drugs.

## Acknowledgements

The authors thank the ELSA team for their hard work and unselfish sharing of survey data.

## Data availability statement

The data used and analyzed in this study are publicly available from the ELSA (https://www.elsa-project.ac.uk). The data that support the findings of this study are available from the corresponding author upon reasonable request.

## Ethics statement

Ethical approval for the ELSA was obtained from the London Multicentre Research Ethics Committee (MREC/01/2/91), and informed consent was gathered from all participants.

## Author contributions

JNH and QMC obtained access to the ELSA data. HBL and QMC conceived and designed the study. JYD performed the statistical analysis. QMC drafted the manuscript and obtained the study funding. YC and JNH carefully revised the manuscript. All authors contributed to the interpretation of data. The corresponding authors attest that all listed authors meet authorship criteria and that no others meeting the criteria have been omitted. All authors read and approved the final manuscript.

## Funding

This work was supported by the National Natural Science Foundation of China (Project No. 82202819, Qingmei Chen), Suzhou Science and Technology Planning Project (SKY2022123, Qingmei Chen), Suzhou Health Youth Backbone Talent “National Tutorial System” Training Project (Qngg2023004, Qingmei Chen).

## Conflict of interest

The authors declare that they have no conflict of interest.

## Notes

### Competing Interest Statement

The authors have declared no competing interest.

### Author Declarations

The London Multicentre Research Ethics Committee (MREC/01/2/91) gave ethical approval for this work.

