## Supplemental Material for "Longitudinal association of remnant cholesterol with cognitive decline vary by lipid-lowering therapy: a population-based cohort study"

**Supplemental Method. Covariates**

We defined wave 2 as the baseline of our study. Hence, covariates were derived from wave 2, unless otherwise indicated. Age data were extracted directly from the database. Sex was categorized as male or female. Education level was classified as NVQ3/GCE A level or NVQ3/GCE A level. In wave 1, the educational level was recorded under the following classification: no qualification, level 1 national vocational qualification (NVQ) or certificate of secondary education, NVQ2 or general certificate of education (GCE) O-level, NVQ3 or GCE A-level, higher qualification but below degree, degree level or higher or NVQ4/5, or others (e.g. refusal). Wave 2 included additional qualifications obtained since wave 1. Qualifications classified as ≥NVQ3/GCE A level included degree/degree level qualification (including higher degree); teaching qualification; nursing qualifications SRN, SCM, SEN, RGN, RM, RHV, Midwife; HNC/HND, BEC/TEC Higher, BTEC Higher/SCOTECH Higher; City and Guilds Full Technological Certificate; SLC/SCE/SUPE at Higher Grade or Certificate of Sixth Year St; SLC Lower; SUPE Lower or Ordinary; School Certificate or Matric; NVQ Level 4/5; or NVQ Level 3/Advanced level GNVQ. The educational data from waves 1 and 2 were combined. Marital status was categorized as married and not married. Current drinking status was grouped into whether or not to drink 1 alcoholic drink per week [1]. Current smoking status was grouped into yes, no, or missing. Body mass index (BMI) was calculated as weight (kg)/height^2^ (m^2^). As some participants missed BMI data in wave 2, we used values from wave 0 and wave 4 as the estimates at wave 2. After the above imputation, there were still 120 participants with missing data on BMI. Physical activity was categorized into inactive (no moderate or vigorous activity weekly) and moderate-vigorous activity (at least once a week). Depressive symptoms were measured with the eight-item version of the Center for Epidemiologic Studies Depression Scale (CESD-8), with a score > 4 indicating depressive symptoms [2]. Hypertension was defined as systolic blood pressure ≥140 mm Hg, diastolic blood pressure ≥ 90 mm Hg, or use of antihypertensive medication. Diabetes was defined as self-reported doctor-diagnosed diabetes, use of anti-diabetic medication, or haemoglobin A1c (HbA1c) ≥ 6.5%. Stroke were defined by self-reported physician diagnoses. In the primary analysis, smoking status was categorized as yes (current smoker), no (non-smoker), and unknown (who were the 0.8% with missing smoking data). Missing BMI and CES-D data were imputed using the mean values of all participants and subsequently converted to categorical values. 42 participants with missing data on physical activity were excluded.

**Table S1. The associations between remnant cholesterol level and rates of cognitive decline according to sex**

| Cognition | Model | Male | |  | Female | | *P* for interaction |
| --- | --- | --- | --- | --- | --- | --- | --- |
|  |  | β (95% CI) | *P* Value |  | β (95% CI) | *P* Value |  |
| Global cognition | 1 | -0.005 (-0.018, 0.007) | 0.411 |  | -0.016 (-0.029, -0.003) | 0.015 | 0.008 |
|  | 2 | -0.005 (-0.018, 0.007) | 0.409 |  | -0.016 (-0.029, -0.003) | 0.015 | 0.009 |
| Semantic fluency | 1 | -0.006 (-0.017, 0.005) | 0.323 |  | -0.009 (-0.019, 0.002) | 0.105 | 0.072 |
|  | 2 | -0.005 (-0.016, 0.006) | 0.345 |  | -0.009 (-0.019, 0.002) | 0.108 | 0.084 |
| Memory | 1 | -0.002 (-0.012, 0.008) | 0.678 |  | -0.010 (-0.020, 0.001) | 0.067 | 0.013 |
|  | 2 | -0.002 (-0.012, 0.008) | 0.699 |  | -0.010 (-0.020, 0.001) | 0.069 | 0.017 |
| Orientation | 1 | -0.005 (-0.020, 0.010) | 0.526 |  | -0.005 (-0.021, 0.010) | 0.479 | 0.308 |
|  | 2 | -0.005 (-0.020, 0.011) | 0.544 |  | -0.006 (-0.021, 0.009) | 0.461 | 0.373 |

Model 1: adjusted for baseline age.

Model 2: further adjusted for low-density lipoprotein cholesterol, BMI, education, marital status, physical activity, current smoking, current drinking, depressive symptoms, hypertension, diabetes, and stroke.

**Table S2. The associations between remnant cholesterol level and rates of cognitive decline according to age**

| Cognition | Model | <65 years | |  | ≥65 years | | *P* for interaction |
| --- | --- | --- | --- | --- | --- | --- | --- |
|  |  | β(95% CI) | *P* Value |  | β(95% CI) | *P* Value |  |
| Global cognition | 1 | -0.011 (-0.020, -0.002) | 0.013 |  | -0.008 (-0.026, 0.009) | 0.365 | 0.954 |
|  | 2 | -0.011 (-0.020, -0.002) | 0.012 |  | -0.008 (-0.025, 0.010) | 0.383 | 0.925 |
| Semantic fluency | 1 | -0.011 (-0.020, -0.002) | 0.021 |  | -0.002 (-0.015, 0.010) | 0.734 | 0.493 |
|  | 2 | -0.011 (-0.020, -0.002) | 0.019 |  | -0.002 (-0.014, 0.011) | 0.805 | 0.529 |
| Memory | 1 | -0.009 (-0.017, -0.001) | 0.036 |  | -0.001 (-0.013, 0.011) | 0.873 | 0.387 |
|  | 2 | -0.009 (-0.018, -0.001) | 0.034 |  | -0.001 (-0.012, 0.011) | 0.928 | 0.398 |
| Orientation | 1 | -0.002 (-0.012, 0.009) | 0.779 |  | -0.008 (-0.029, 0.014) | 0.477 | 0.398 |
|  | 2 | -0.002 (-0.012, 0.009) | 0.777 |  | -0.007 (-0.029, 0.014) | 0.497 | 0.426 |

Model 1: adjusted for sex.

Model 2: further adjusted for low-density lipoprotein cholesterol, BMI, education, marital status, physical activity, current smoking, current drinking, depressive symptoms, hypertension, diabetes, and stroke.

**Table S3. The associations between remnant cholesterol level and rates of cognitive decline according to education levels**

| Cognition | Model | < NVQ3/GCE A level | |  | ≥NVQ3/GCE A level | | *P* for interaction |
| --- | --- | --- | --- | --- | --- | --- | --- |
|  |  | β(95% CI) | *P* Value |  | β(95% CI) | *P* Value |  |
| Global cognition | 1 | -0.006 (-0.018, 0.005) | 0.304 |  | -0.013 (-0.027, 0.001) | 0.066 | 0.627 |
|  | 2 | -0.006 (-0.017, 0.006) | 0.320 |  | -0.013 (-0.027, 0.001) | 0.060 | 0.563 |
| Semantic fluency | 1 | -0.007 (-0.016, 0.002) | 0.129 |  | -0.007 (-0.021, 0.006) | 0.298 | 0.949 |
|  | 2 | -0.007 (-0.016, 0.002) | 0.134 |  | -0.008 (-0.021, 0.006) | 0.279 | 0.861 |
| Memory | 1 | -0.011 (-0.020, -0.002) | 0.018 |  | 0.006 (-0.006, 0.018) | 0.329 | 0.053 |
|  | 2 | -0.011 (-0.019, -0.002) | 0.019 |  | 0.006 (-0.006, 0.018) | 0.351 | 0.068 |
| Orientation | 1 | 0.004 (-0.010, 0.019) | 0.543 |  | -0.016 (-0.031, 0.000) | 0.048 | 0.579 |
|  | 2 | 0.004 (-0.010, 0.019) | 0.543 |  | -0.016 (-0.032, 0.000) | 0.044 | 0.520 |

Model 1: adjusted for sex and age.

Model 2: further adjusted for low-density lipoprotein cholesterol, BMI, marital status, physical activity, current smoking, current drinking, depressive symptoms, hypertension, diabetes, and stroke.

**Table S4. Associations between baseline remnant cholesterol levels and rates of cognitive decline**

**after excluding participants who experienced stroke during the follow-up**

|  | Model | β (95% CI) | *P* Value |
| --- | --- | --- | --- |
| Non-lipid-lowering group |  |  |  |
| Global cognition | 1 | -0.019 (-0.031, -0.006) | 0.003 |
|  | 2 | -0.018 (-0.031, -0.006) | 0.003 |
| Semantic fluency | 1 | -0.006 (0.000, 0.004) | 0.247 |
|  | 2 | -0.006 (-0.017, 0.005) | 0.262 |
| Memory | 1 | -0.009 (-0.019, 0.001) | 0.089 |
|  | 2 | -0.008 (-0.018, 0.002) | 0.098 |
| Orientation | 1 | -0.015 (-0.030, 0.000) | 0.047 |
|  | 2 | -0.015 (-0.030, 0.000) | 0.046 |
| Lipid-lowering group |  |  |  |
| Global cognition | 1 | 0.002 (-0.012, 0.015) | 0.781 |
|  | 2 | 0.001 (-0.012, 0.015) | 0.846 |
| Semantic fluency | 1 | -0.006 (-0.018, 0.006) | 0.307 |
|  | 2 | -0.007 (-0.019, 0.005) | 0.279 |
| Memory | 1 | -0.002 (-0.013, 0.009) | 0.686 |
|  | 2 | -0.003 (-0.014, 0.009) | 0.642 |
| Orientation | 1 | 0.014 (-0.003, 0.03) | 0.108 |
|  | 2 | 0.014 (-0.003, 0.03) | 0.112 |

Model 1: adjusted for sex and age.

Model 2: further adjusted for low-density lipoprotein cholesterol, BMI, education levels, marital status, physical activity, current smoking, current drinking, depressive symptoms, hypertension, diabetes, and stroke.

**Table S5. Associations between baseline remnant cholesterol levels and rates of cognitive decline**

**after restricting participants to those who attended all cognitive assessment**

|  | Model | β (95% CI) | *P* Value |
| --- | --- | --- | --- |
| Non-lipid-lowering group |  |  |  |
| Global cognition | 1 | -0.015 (-0.028, -0.002) | 0.022 |
|  | 2 | -0.014 (-0.027, -0.002) | 0.026 |
| Semantic fluency | 1 | -0.006 (-0.019, 0.006) | 0.306 |
|  | 2 | -0.006 (-0.018, 0.006) | 0.339 |
| Memory | 1 | -0.012 (-0.023, 0.000) | 0.044 |
|  | 2 | -0.011 (-0.023, 0.000) | 0.049 |
| Orientation | 1 | -0.012 (-0.027, 0.004) | 0.132 |
|  | 2 | -0.012 (-0.027, 0.004) | 0.136 |
| Lipid-lowering group |  |  |  |
| Global cognition | 1 | 0.004 (-0.009, 0.018) | 0.542 |
|  | 2 | 0.004 (-0.010, 0.018) | 0.579 |
| Semantic fluency | 1 | -0.003 (-0.016, 0.010) | 0.631 |
|  | 2 | -0.003 (-0.017, 0.010) | 0.605 |
| Memory | 1 | 0.002 (-0.010, 0.013) | 0.774 |
|  | 2 | 0.002 (-0.010, 0.014) | 0.735 |
| Orientation | 1 | 0.009 (-0.008, 0.026) | 0.295 |
|  | 2 | 0.010 (-0.007, 0.028) | 0.242 |

Model 1: adjusted for sex and age.

Model 2: further adjusted for low-density lipoprotein cholesterol, BMI, education levels, marital status, physical activity, current smoking, current drinking, depressive symptoms, hypertension, diabetes, and stroke.

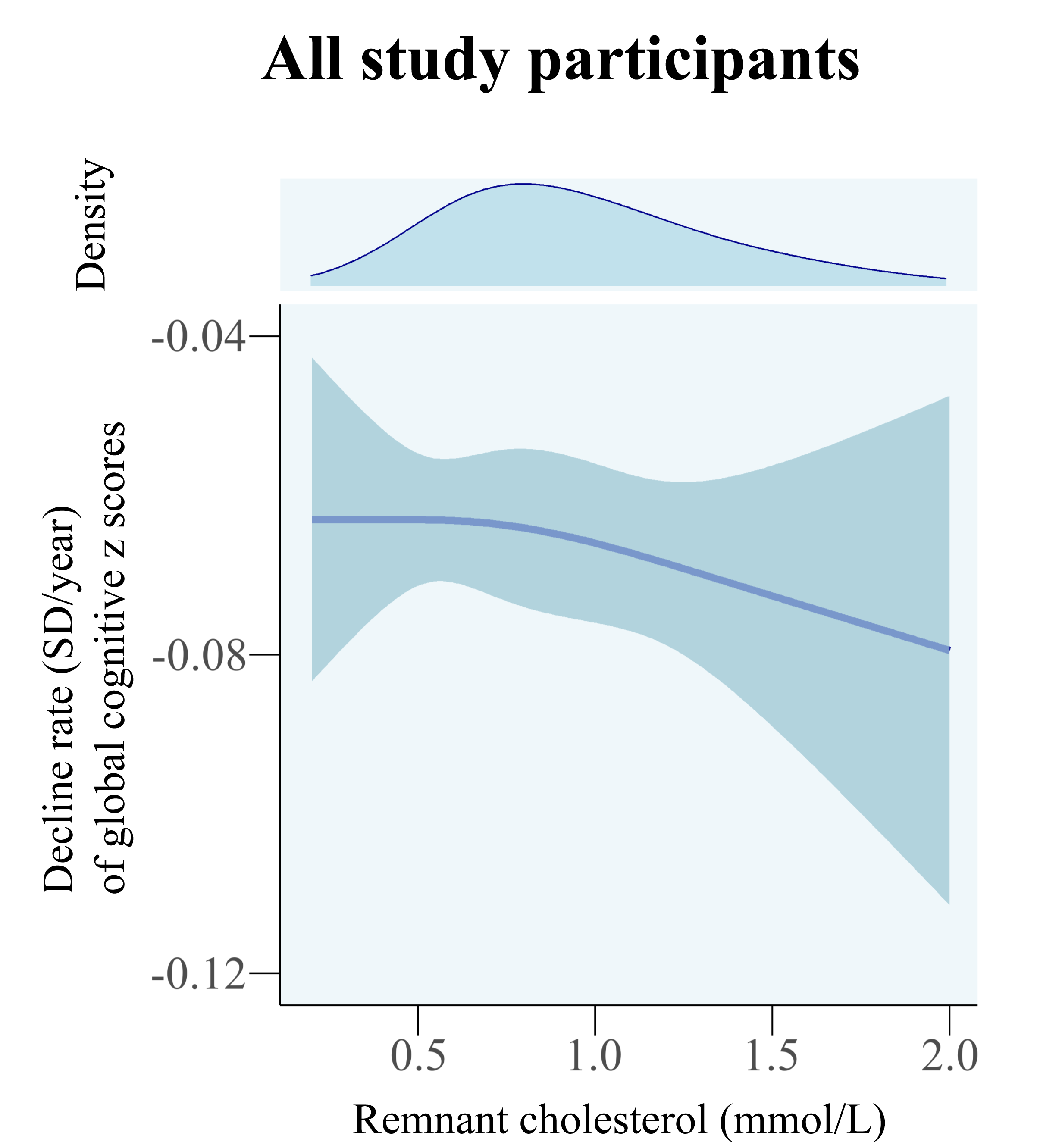

**Figure S1. Dose-response associations of remnant cholesterol levels at baseline with rate of global cognitive decline over the follow-up.**

Note, based on model fit statistics of Akaike Information Criterion, three knots were placed at the 10^th^, 50^th^, and 90^th^ percentiles of remnant cholesterol values. Models were adjusted for age, sex, low-density lipoprotein cholesterol, body mass index, education, marital status, physical activity, current smoking, current drinking, depressive symptoms, hypertension, diabetes, and stroke.
